# Driving uptake of missed routine vaccines in adolescent and adult migrants: a prospective observational mixed-methods pilot study of catch-up vaccination in UK general practice

**DOI:** 10.1101/2023.07.03.23292165

**Authors:** Alison F Crawshaw, Lucy P Goldsmith, Anna Deal, Jessica Carter, Felicity Knights, Farah Seedat, Karen Lau, Sally E Hayward, Joanna Yong, Desiree Fyle, Nathaniel Aspray, Michiyo Iwami, Yusuf Ciftci, Fatima Wurie, Azeem Majeed, Alice S Forster, Sally Hargreaves

## Abstract

**Background:** Migrants in Europe may be vulnerable to vaccine preventable diseases (VPDs) because of missed routine vaccines in childhood in their country of origin and marginalisation from health and vaccine systems. To align with European schedules, migrants should be offered catch-up vaccinations, considering MMR, Td/IPV, and age-appropriate MenACWY and HPV. However, awareness and implementation of catch-up guidelines by primary care staff in the UK is considered to be poor, and there is a lack of research on effective approaches to strengthen the primary-care pathway.

**Methods:** We conducted a prospective observational mixed-methods pilot study ‘Vacc on Track’ (May 2021-September 2022) to better understand and define new care pathways to increase catch-up vaccination for adolescent and adult migrants presenting to primary care (≥16 years, born outside Western Europe, North America, Australia, or New Zealand) in two London boroughs. We designed a standardised data collection tool to assess rates of under-vaccination in migrant populations and previous VPDs, which then prompted a referral to practice nurses to deliver catch-up vaccination for those with uncertain or incomplete immunisation status, following UK guidelines. We explored views of practice staff on delivering catch-up vaccination to migrant populations through focus group discussions and engaged migrants in in-depth interviews around approaches to catch-up vaccination. Data were analysed in STATA12 and Microsoft Excel.

**Results:** We recruited 57 migrant participants (mean age 41 [SD 7.2] years; 62% female; mean 11.3 [SD 9.1] years in UK) from 18 countries, with minimum 6 months’ follow-up. We did 3 focus groups with 30 practice staff and 39 qualitative in-depth interviews with migrants. Nearly all migrant participants required catch-up vaccination for MMR (86%) and Td/IPV (88%) and most reported not having been previously engaged in UK primary care around catch-up vaccination. 12 (55%) of 22 participants in Site 1 reported a past VPD, including measles and rubella. 53 (93%) of participants were referred for catch-up vaccination. However, although 43 (81%) had at least one dose (at follow-up) of a required vaccine, only 6 (12%) referred for Td/IPV and 33 (64%) of those referred for MMR had completed their required course and vaccination pathway at follow-up, suggesting there were a range of personal and environmental obstacles to migrants accessing vaccinations and all multiple doses of vaccines that need to be better considered. Staff identified seven barriers to delivering catch-up vaccines to migrants, including limited time for appointments and follow-up, language and literacy barriers when taking histories and to encourage vaccination, lack of staff knowledge of current guidelines, inadequate engagement routes, and the absence of primary care targets or incentives.

**Conclusions:** Our findings suggest adolescent and adult migrants are an under-vaccinated group and would benefit from being offered catch-up vaccination on arrival to the UK. Primary care is an important setting to deliver catch-up vaccination, but effective pathways are currently lacking, and improving vaccine coverage for key routine vaccines across a broader range of migrant groups will require designated staff champions, training, awareness-raising and financial incentives. Novel ways to deliver vaccinations in community settings should be explored, along with co-designing community-based interventions to raise awareness among these populations of the benefits of life-course immunisation.

## Introduction

Adult and adolescent migrants to Europe may be under-immunised and vulnerable to vaccine preventable diseases (VPDs) due to missing routine vaccinations in childhood and marginalisation from health systems and should be aligned with European immunisation schedules (1–4). Indeed, migrant populations have been involved in outbreaks of VPDs, including measles, in Europe (5), and immunisation gaps and disparities may now be widened following the COVID-19 pandemic, which had a detrimental effect on the delivery of routine immunisation programmes globally (6). In a recent analysis of 125,526 refugees in an IOM resettlement programme to the UK, for example, only 11% of refugees were fully aligned with the UK schedule for polio, 34% for measles, and 5% for diphtheria and tetanus, with adults more likely than children to be under-immunised, suggesting that refugees, and the broader migrant population in Europe, could benefit from being offered catch-up vaccinations in UK primary care on arrival (3). Adolescent and adult migrants are, however, rarely considered in vaccination programmes on arrival to European countries (7), with the focus predominantly on very young children and specific groups (8). Despite clear guidelines for offering catch-up vaccinations to migrants in the UK, awareness and implementation are low in practice (9). Better understanding of the barriers and facilitators to catch-up vaccination in mobile and migrant populations is urgently needed to reach immunisation coverage targets and ensure equitable access to vaccines (10).

The reasons why some migrants are at risk of under-immunisation for routine vaccinations are well documented (11–15), and include a range of cultural, socio-structural, political, economic, and behavioural factors, including language barriers and a lack of specific procedures to engage older age groups in catch-up vaccination. Migrants’ countries of origin often have differing immunisation schedules, reduced availability of vaccines, poor health system infrastructure or fragmented delivery systems so these populations in Europe and the UK may have missed vaccines, doses, and boosters and not have been offered newer vaccines such as HPV that were not available in their host countries. While in host and destination countries, migrants may be reluctant to engage with vaccination services due to mistrust of healthcare systems, public health or immigration authorities and governments, forms of racism and discrimination, physical barriers to access, and specific beliefs or concerns about vaccines. Too often, the onus is on migrants to engage better with health systems and services, rather than on developing more equitable and inclusive approaches and policies to meaningfully engage with these groups. Unlike migrant children, who are typically caught up for missed childhood vaccines through the school system, adult and adolescent migrants are particularly likely to remain unvaccinated during and after migration due to weak systems to facilitate checking vaccination history and offering catch-up vaccinations and inconsistent implementation and interpretation of guidance.

Various frameworks exist to drive improvement vaccination coverage for VPDs and ensure equitable access and uptake of vaccinations by migrant populations (10, 16–19). The World Health Organization’s (WHO) new Immunization Agenda 2030 (IA2030) sets out strategic priorities to strengthen immunisation within primary care, increase equitable access to vaccination for vulnerable populations, and integrate catch-up vaccination for missed vaccines/doses throughout the life-course. It recommends that all countries have a catch-up vaccination policy and schedule in place, and it provides specific guidance for countries (10), as does the European Centre for Disease Prevention and Control (ECDC) (20) for countries in Europe. In the UK, the UK Health Security Agency (UKHSA) provides specific guidance on the vaccination of individuals with uncertain or incomplete immunisation status (21), which is relevant to many migrants, most of whom arrive with no vaccine records or who have no recorded history of childhood vaccines on primary care systems. It states that unless there is a documented or reliable verbal vaccine history, individuals should be assumed to be unimmunised, and a full course of immunisations planned (21). However, implementation of this guidance in primary care is poor, with adult migrants typically excluded from catch-up vaccination initiatives due to lack of pathways in place to engage these individuals on presentation to primary care and lack of knowledge among front-line staff on effective approaches (9) . Moreover, data on migrant status (foreign born, country of origin) are not routinely coded into electronic patient records in UK primary care, making it difficult to understand and address the vaccination needs of specific migrant groups, understand levels of under-immunisation, and gaps in provision around catch-up vaccination.

We therefore did a prospective, observational mixed-methods pilot study (Vacc on Track), to better understand and define new care pathways using a standardised data collection tool to engage and facilitate the inclusion of adolescent and adult migrants in catch-up vaccination initiatives in UK primary care, ahead of a potential larger-scale trial.

**Figure 1.**
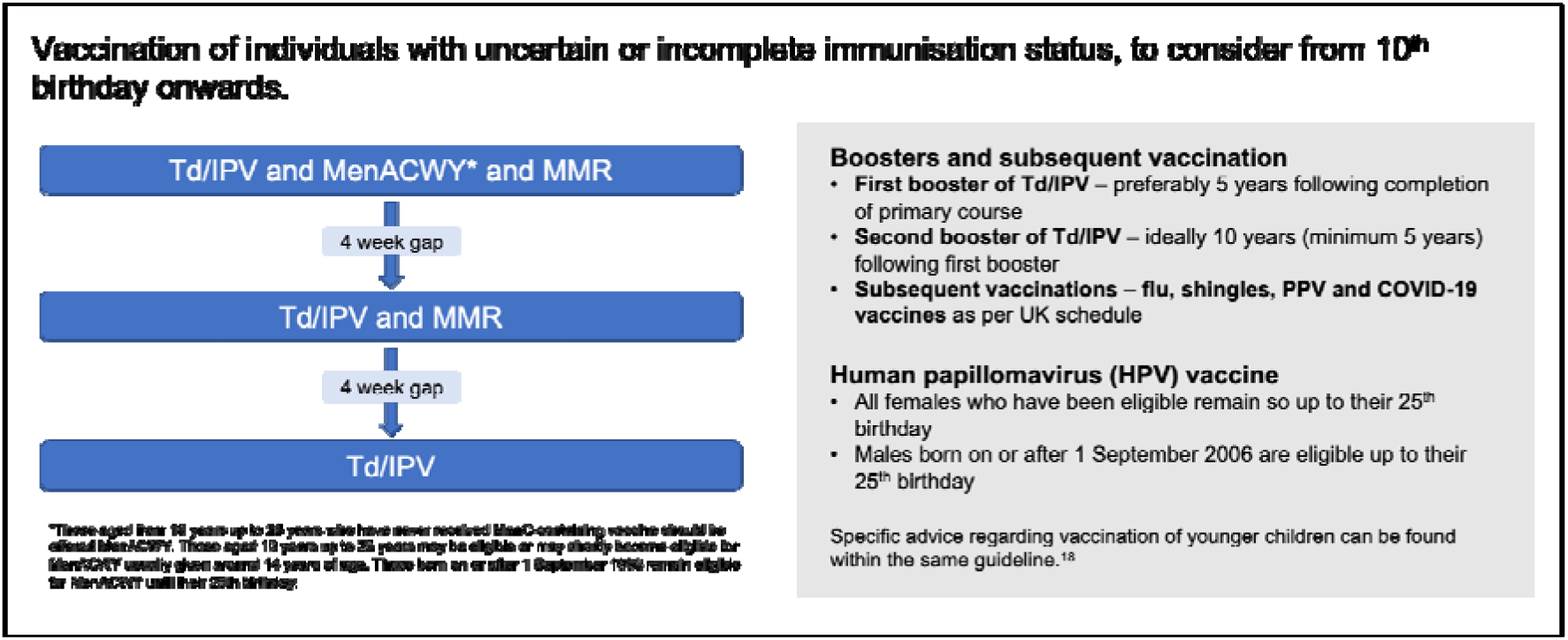
Vaccination of adolescent and adult individuals with uncertain or incomplete immunisation status. Reproduced from (21). Guidance states unless there is a documented or reliable verbal vaccine history, individuals should be assumed to be unimmunised, and a full course of immunisations planned. MMR=measles, mumps, rubella; Td/IPV= tetanus, diphtheria, polio; HPV= human papillomavirus vaccine; PPV=pneumococcal vaccine; MenACWY= meningococcal conjugate vaccine. Guidance for other age groups (<10 years) is provided in the original source.

## Methods

### Study design and setting

We conducted a prospective, observational mixed-methods pilot study from May 2021-September 2022 in seven GP practices across two urban London boroughs. Boroughs were selected for their high proportion of migrant residents (estimated to be approximately half, according to 2021 Census data (22)). Migrants were defined as individuals who were born outside of Western Europe, North America, Australia, and New Zealand. We designed a standardised data collection tool (Figure 2) to collect quantitative data on rates of under-immunisation and history of VPDs that then prompted referrals to practice nurses for catch-up vaccinations in adult and adolescent migrants who required additional vaccines and boosters as per UK guidelines. Catch-up vaccinations should be part of routine care and include MMR, Td/IPV, HPV (aged 11-25 years) and MenACWY (aged 10-25 years) vaccines for those with uncertain or incomplete immunisation status, according to the UK catch-up vaccination guidelines (21).

**Figure 2.**
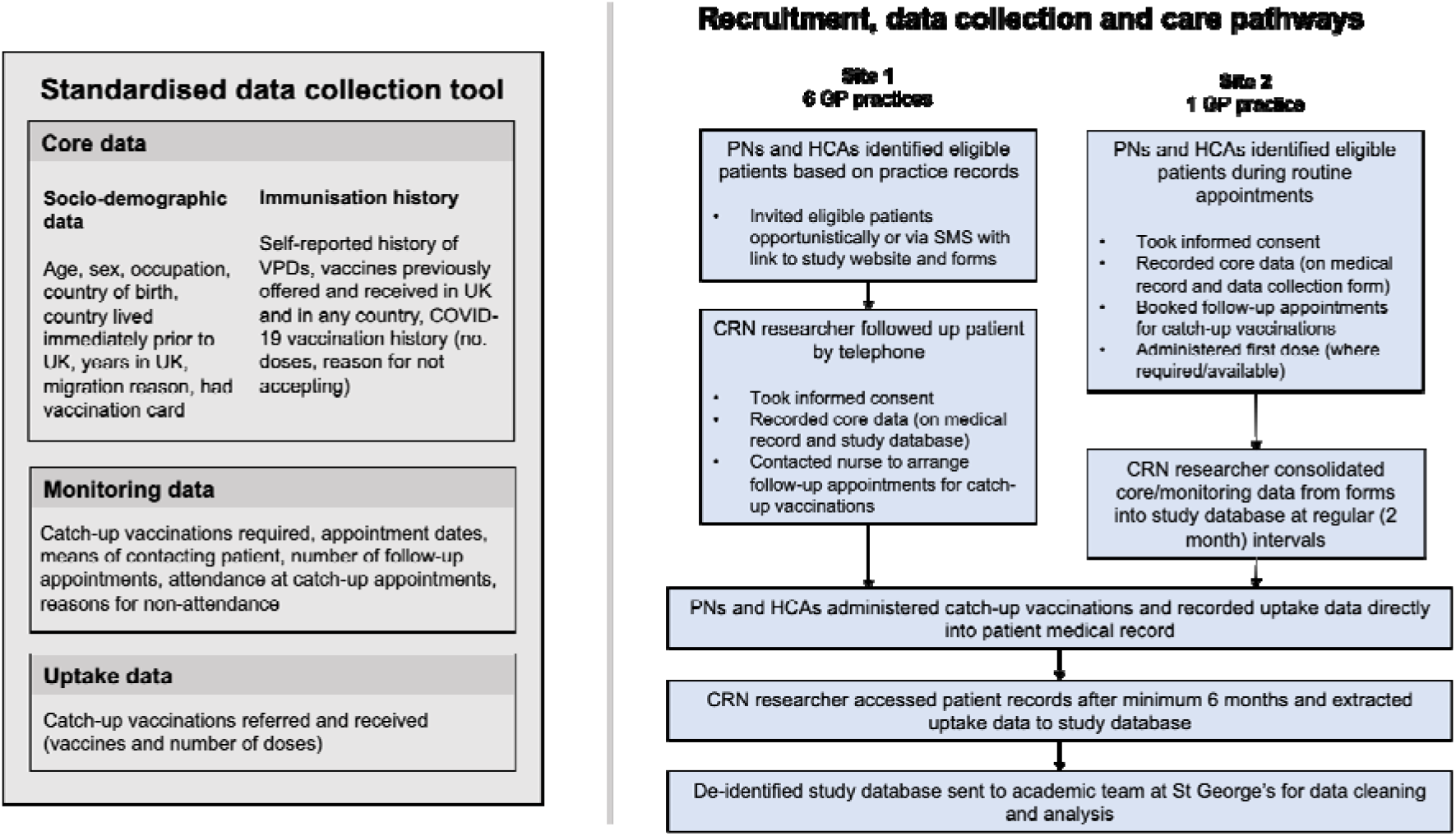
Figure showing standardised data collection tool (left) and care pathways implemented in study sites 1 & 2 (right). VPDs = vaccine-preventable diseases, PN = practice nurse, HCA = healthcare assistant, CRN = clinical research network.

PICOTS criteria for the study are shown in Box 1. In additional to collecting quantitative data from migrant patients, we explored the views of practice staff on catch-up vaccination and current guidance, including barriers to implementation, suggestions, and areas for improvement and support, through focus group discussions (FGDs). During the study, we also decided to conduct a key informant interview to explore examples of good practice from the most successful (in terms of recruitment and uptake) participating practice. We also carried out in-depth interviews with a diverse range of recently arrived migrants to explore views and concerns around catch-up vaccination after arrival in the UK (reported elsewhere). The study tool, recruitment and data collection pathways are shown in Figure 2. The reporting of this study follows STROBE guidelines (23).

**Box 1.**
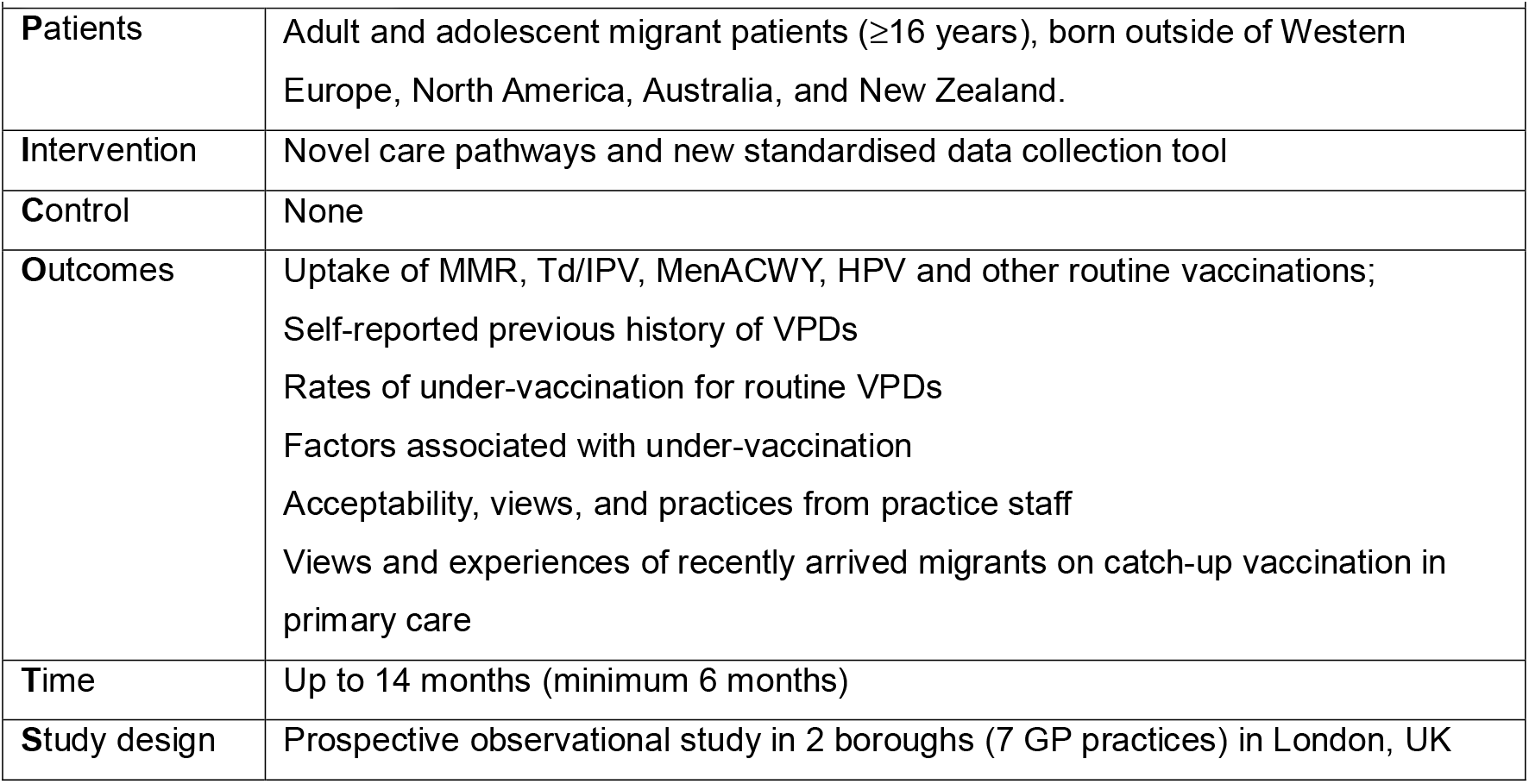
PICOTS criteria.

### Ethics and PPIE

This study received ethics approval from the NHS Health Research Authority Yorkshire and Humber – South Yorkshire Research Ethics Committee (20/YH/0342) on 18th December 2020. The qualitative in-depth interview study with migrants received ethics approval from the St George’s, University of London Research Ethics Committee (REC reference: 2020.0058). Migrants with lived experience of the UK immigration and healthcare systems were involved in the design of this study through our National Institute for Health and Care Research (NIHR) funded Patient and Public Involvement and Engagement (PPIE) Project Advisory Board and were compensated for their time and contributions.

### Participants, recruitment, and informed consent process

The study was conducted with support from the NHS North Central London Research Network (NoCLoR) and the North Thames Clinical Research Network (CRN). GP practices in areas with a high proportion of migrant residents were purposively invited to join the study. We aimed to recruit up to 10 GP practices across two boroughs (Barnet and Tower Hamlets) in North and East London (referred to henceforth as Site 1 & 2), with a target sample size of 100 participants. Both boroughs rank in the top 50% of most deprived local authorities in England (24), although Tower Hamlets ranks as significantly more deprived than Barnet.

We held site initiation visits with all practice sites, inviting GPs, practice managers, HCAs, and nurses involved in immunisation, where we summarised current UK primary care catch-up vaccination guidelines, and showed the team the approach to identifying migrants, facilitating data collection via the standardised data collection tool, and communicated care pathways for under-immunised migrants that were identified.

Patients registered at participating practices were eligible for the study if they were a) aged 16 years or older, b) born outside of the UK (excluding North America, Australia, New Zealand, or Western Europe, as defined by the UN maximal definition of Western Europe (25)), and c) capable of giving informed consent. Recruitment procedures differed between the two sites (see Figure 2).

At Site 1, which comprised 6 GP practices, clinical practice staff were originally going to recruit and consent patients. However, the recruitment pathway was modified as clinical staff were under intense pressure from the COVID-19 pandemic, so the CRN led the recruitment and consenting process. Practice nurses and healthcare assistants (HCAs) first identified patients who met the eligibility criteria, filtering patient records by ethnicity or notes on migrant status (where recorded) to identify those potentially eligible and sent an SMS/text message with a link to the study website, from which patients could download the study documents (participant information sheet [PIS], consent form, and leaflets about catch-up vaccination and HPV vaccination, all available in the six dominant local languages, which were Arabic, Farsi, Pashtu, Romanian, Urdu, English). A researcher at the CRN (DF) then followed up with patients by a telephone call enquiring whether they would like to join the study and to take informed consent. Practice nurses also mentioned the study opportunistically to patients during routine appointments, who would then be referred to the CRN researcher (DF) for consent. At Site 2, which comprised one GP practice, the practice nurses and HCAs invited and consented participants to the study opportunistically during routine appointments, as per the original recruitment pathway. Participants were given hard copies of the study documents and given the opportunity to ask questions and decide whether they wanted to participate. We gave practice and CRN staff a copy of a form detailing the names of common childhood vaccines in multiple languages, to support taking vaccine history during appointments (see supplementary files). Telephone interpreters (via Language Line) were available on request at both sites during recruitment and data collection.

### Data collection and care pathway for catch-up vaccination

The standardised data collection tool was developed by the study team in Microsoft Excel and facilitated the recording of specific immunisation history and socio-demographic data (‘core data’), alongside monitoring and vaccine uptake data from patients (Figure 2). From these data, we documented participants’ rates of under-vaccination for MMR, Td/IPV, and other key vaccines in the UK routine immunisation schedule, history of VPDs, and uptake rates of MMR, Td/IPV, MenACWY, and HPV vaccines following referral to the practice nurse. We also explored socio-demographic factors associated with under-vaccination in the study population. Immunisation history was based on self-reporting or vaccination records (via the primary care computer system or hand-held vaccination cards) where available. Monitoring data included catch-up vaccinations required, appointment dates, means of contacting patients and number of follow-ups, attendance at catch-up vaccination appointments and reasons for non-attendance. Uptake data included final catch-up vaccines and doses received through the study.

Data collection and care pathways and procedures differed between sites and are outlined in Figure 2. In site 1, the CRN researcher collected core data via telephone call with the participant, which were recorded in the patient’s electronic medical record and on a password-protected study database. The CRN researcher determined the participant’s need for catch-up vaccinations based on the study training and the UK catch-up vaccination guidelines (21) and, if acceptable to the participant, this data prompted contact with the practice nurse to arrange an appointment. Once the CRN staff had facilitated an appointment for first doses, they then left practice nurses to follow-up patients for subsequent doses as per routine care. Subsequent catch-up vaccination doses (uptake data) were recorded by practice staff in the patient’s medical record at the time of administration and these data were later extracted by the CRN researcher. In Site 2, the practice nurse collected core data (recorded in the patient’s medical record) during face-to-face appointments and this prompted a referral for a booking with the immunisation team at the practice for necessary follow-up appointments for catch-up vaccinations and subsequent doses.

### Data management, follow-up, and statistical analysis

We aimed to follow up patients for a minimum of 6 months at both sites to allow for all doses to be given (Td/IPV is 3 doses, with a 4 week gap in between each, MMR is 2 doses with a 4 week gap); Figure 1). At the end of follow-up, the anonymised study data (core, monitoring and uptake data) were extracted from participants’ electronic medical records by the CRN researcher (at Site 1) or the practice manager at Site 2 and securely transferred to the CRN researcher, who updated the aggregate study database. A de-identified, anonymised version was then transferred securely to the study team at St George’s for data cleaning and analysis.

Data cleaning and analyses were done using STATA 12. All tests were two-tailed and p values less than 0.05 were regarded as significant. We used descriptive statistics to describe the socio-demographic characteristics, vaccination history, VPD history and catch-up vaccine uptake of participants. We summarised continuous data with mean and standard deviation (SD) and described categorical responses using frequency and percentage. Comparisons between categorical variables were calculated using Pearson’s Chi-squared test and comparisons between continuous variables were calculated using unpaired t-tests.

Bivariable and multivariable logistic regression analyses were used to look for factors associated with being un-vaccinated (received zero doses) or under-vaccinated (received at least 1 dose, but not full schedule) for key vaccines at the time of study enrolment. Outcomes included un-vaccinated for MMR vaccine; un-vaccinated for Td/IPV vaccine; un-vaccinated for MMR vaccine *and* Td/IPV vaccine; unvaccinated for any polio – combined or single vaccines; unvaccinated for any measles – combined or single vaccines; and under-vaccinated for MMR vaccine or Td/IPV vaccine. Explanatory variables were age, sex, birth region, region lived prior to the UK, years in the UK, and study site (migration reason and occupation were only recorded in Site 1 and were therefore not included in the regression analyses). Multivariable models were built in a forward, stepwise fashion. Age, sex, and birth region were adjusted for in each multivariable model; certain variables were removed from the final model to reduce collinearity. Unadjusted and adjusted odds ratios were calculated using generalised estimating equations logistic regression.

### Qualitative component

Our qualitative component included focus group discussions (FGDs) and a key informant interview conducted with practice staff from participating practices and in-depth interviews conducted with recently arrived migrants. The in-depth interviews with migrants (methods and findings) are reported in full elsewhere. We did three FGDs which were scheduled as part of routine practice meetings conducted on Microsoft Teams (most convenient for participants). Participants were practice nurses, healthcare assistants (HCAs), and practice managers (roles involved in vaccination delivery/scheduling) from the participating practices. A key informant interview was conducted with two staff from Site 2 who had not participated in FGDs. For the FGDs, all staff received information about the study and how their data would be used in advance, which was reiterated at the start of the meeting, and staff were able to make an informed decision about their participation. Participants were asked to imply consent by remaining on the call, which was considered appropriate because the topic was low risk, not audio recorded, and anonymised summary feedback (broad views) was collected.

Key informant interview participants received a PIS and provided written informed consent prior to participating. Both the FGD and key informant interview followed a semi-structured topic guide, which explored participants’ experiences of implementing the study, current barriers and challenges to delivering catch-up vaccinations, and suggestions for improving the tool, care pathways, and engaging migrant patients/promoting catch-up vaccination among these groups. Broad views and selected short-hand quotations (non-attributable) were collected during FGDs in the form of hand-written and typed notes (by SH and LPG). The key informant interview was conducted by AFC with two staff participants in a private room, audio-recorded and transcribed verbatim by a professional transcription service.

Qualitative data were analysed deductively using a flexible and rapid thematic analysis and evaluation approach (26). Notes from the FGDs were reflected on and discussed afterwards by AFC, SH and LPG, and AFC then independently coded and grouped the findings into broad barrier and facilitator concepts using a matrix method (by hand and in Microsoft Excel). The data in the matrix were corroborated and discussed again by the three researchers, to ensure rigour and coding reliability.

The same approach was used to analyse the transcript of the key informant interviews. Triangulation occurred when the qualitative and quantitative data were combined but also by the interaction between the three researchers during data collection and analysis, and through the contributions of their own perceptions, beliefs and academic disciplines to the collection and interpretation of data.

## Results

We recruited 57 migrant patient participants to the study between May 2020-September 2021 from seven GP practices. Site 1 comprised 22 participants in six practices in Barnet, North London. Site 2 comprised 35 participants in one practice in Tower Hamlets, East London. Participants were followed up for up to 14 months in Site 1 (median: 12 months, 1 participant followed up for 2 months only due to late recruitment) and for 6 months in Site 2. We conducted 3 FGDs with a total of 30 practice staff (practice nurses, HCAs, and practice managers), one key informant interview with two practice staff (lead practice nurse and assistant practice manager) and 39 in-depth interviews with migrants (reported elsewhere).

### Socio-demographic description of migrant participants

The mean age of the combined study population was 41 years (SD: 7.2 years); 62% were female. Participants had spent a mean 11.3 years (SD: 9.1 years) in the UK at the time of recruitment and came from 18 different countries of birth across Eastern Europe, Africa, Latin America and the Caribbean, and Asia, although the majority (75%) were born in Asia. 16% of participants had a vaccination card. Half (50%) of participants from Site 1 migrated to the UK for economic reasons, with smaller proportions reporting forced migration, study, or joining/accompanying family. Two-thirds (64%) of participants from Site 1 were currently working in higher-skilled jobs (based on ONS Labour Force Survey categories (27)). Data on migration reasons and occupation were not collected from Site 2. Demographic distributions did not differ statistically significantly between sites for age, sex, or years in UK, but did differ by birth region (p=0.01) and possession of vaccination card (p=0.009) (Table S6). Demographic characteristics are shown in Supplementary Table S1.

[Link to Supplementary Files – Table S1]

### Under-vaccination for routine and selective/travel vaccines at study outset

A high proportion of our study population were incompletely vaccinated or unvaccinated according to UK immunisation guidelines (21) for several routine vaccines (Figure 3; Table S2). Specifically, 86% (49) of participants were incompletely vaccinated or unvaccinated for measles, mumps, and rubella vaccines and 88% (50) for diphtheria, tetanus, and polio vaccines (Table 1). Forty-seven (82%) participants had never received any measles-containing vaccine (single or combined vaccines) and 27 (47%) participants had never received any polio-containing vaccine.

**Table 1.**
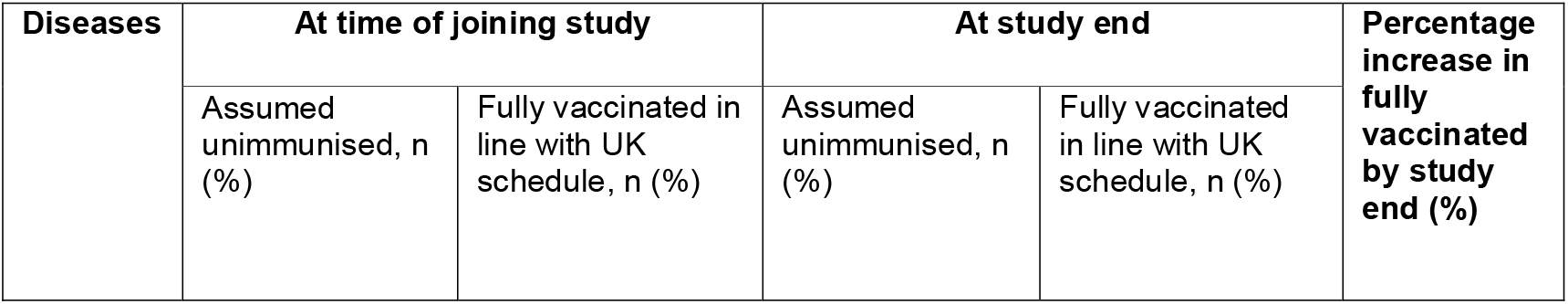
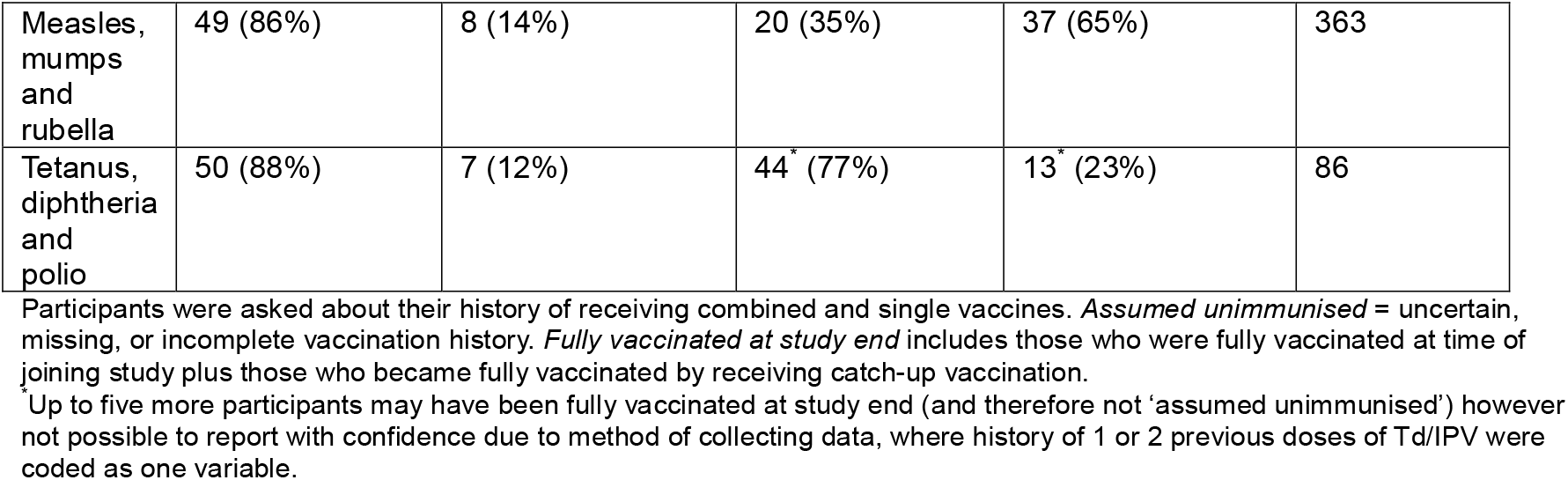
Participants assumed un-immunised and fully vaccinated in line with UK schedule for measles, mumps, rubella, tetanus, diphtheria, and polio at time of joining study and at study end (n=57).

Among selective vaccinations (offered according to criteria such as age), 14 (25%) participants had received an HPV vaccination in any country, and 11 (19%) participants had received MenACWY vaccination in any country (Table S2); neither of the 2 (4%) participants who were currently eligible for these vaccinations based on age had received either vaccine at the time of joining the study.

In addition, 86% reported never having received a TB/BCG vaccine, and 91% had never received a hepatitis B vaccine. Migrants reported high levels of COVID-19 vaccination: most (53, 93%) had received at least one dose of the COVID-19 vaccine (54% had received 3 doses, 35% – 2 doses; 4% – 1 dose) (Table S7).

[Link to supplementary files – Tables S2 and S7]

**Figure 3.**
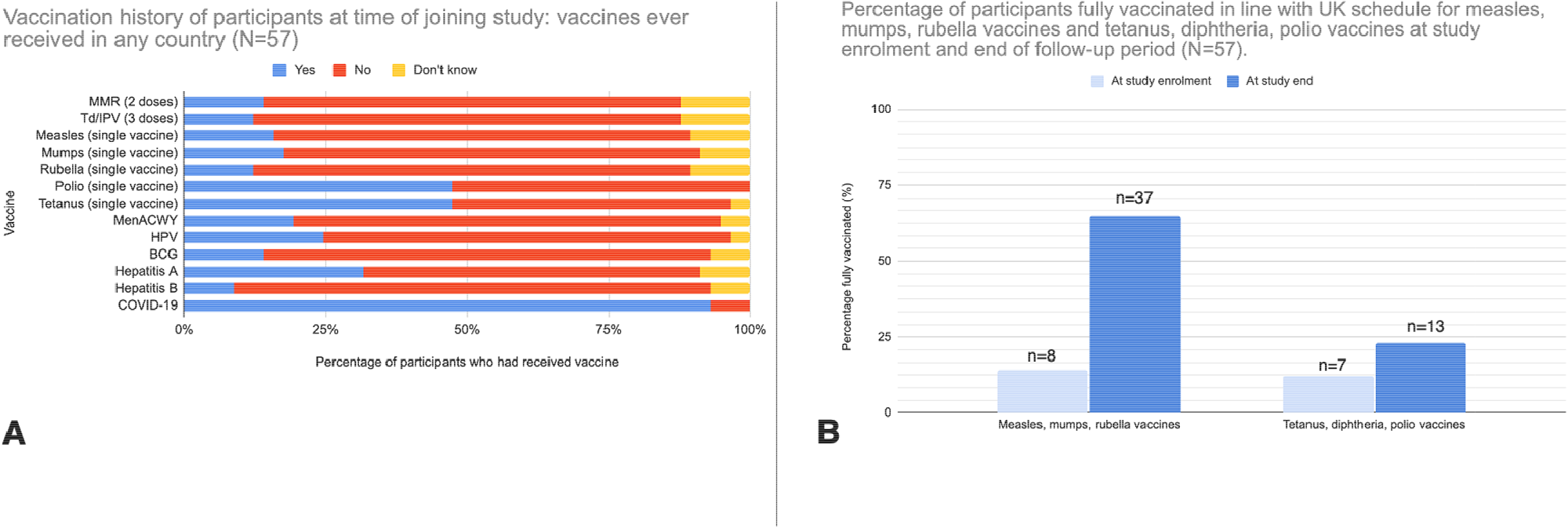
**A:** Participants’ vaccination history at time of joining the study {N=57), self-reported in response to the question *’Have you ever received [vaccine name]? If yes, how many doses did you receive?’* and recorded as yes, no, or don’t know. Uptake of complete dosages shown for MMR (2 doses) and Td/lPV (3 doses); history of receiving at least one dose shown for all other vaccines. **B:** Percentages of participants fully vaccinated in line with UK schedule for measles, mumps, rubella vaccines and tetanus, diphtheria, pertussis vaccines at study enrolment and end of follow-up period (N=57).

### Factors associated with under-vaccination for key vaccines

There were no statistically significant (p<0.05) associations on bivariable or multivariable analysis with any of our outcomes except for being un-vaccinated for polio (0 doses of combined or single vaccine) (Table S5). On multivariable analysis, after adjusting for age, sex and birth region, the odds of being unvaccinated for polio (0 doses) significantly increased by 21% for every 1-year increase in age (aOR: 1.21 [1.01-1.47] and significantly decreased by 25% for every additional year spent in the UK ([aOR: 0.73 [0.60-0.94]).

[Link to Supplementary Files – Table S5]

### Vaccinations offered and received in the UK (prior to study participation)

All participants reported having been offered vaccines in the UK, and 88% said that they had received something. The most offered vaccines were COVID-19 vaccine (100%), influenza vaccine (51%), hepatitis A (21%) and typhoid vaccine (18%). Less than 10% of participants were offered any of MMR, Td/IPV, MenACWY, HPV, which are catch-up vaccines as per UK guidelines (Table S2).

[Link to Supplementary Files – Table S2]

### History of VPDs in presenting migrants

History of VPDs was collected from Site 1 participants (and both sites for COVID-19 disease) and is shown in Table S3. Half of patients from Site 1 (12 [55%] of 22 participants) recalled having a VPD (exact timing unknown and not including COVID-19), and 3 (14%) participants reported having had two or more VPDs (not including COVID-19). Reported VPDs included measles (n=2), rubella (n=1), active TB (n=1), bacterial meningitis (n=1), pertussis (n=2), hepatitis A (n=1), hepatitis B (n=1), and HPV (n=5) (Table 2). 50 (88%) of 57 participants reported having had COVID-19 disease (Table S3).

[Link to Supplementary Files – Table S3]

### Uptake of catch-up vaccinations

The aggregated data show that 53 (93%) participants were referred for catch-up vaccinations as part of the study (Table S4). Three quarters (43, 75%) received at least one dose of any catch-up vaccination. The most common reason for not receiving at least one dose was loss-to-follow-up from not responding to the invitation (5/10, 50%). Table 1 and Figure 3B show the proportion of participants who were fully vaccinated in line with the UK schedule for measles, mumps, rubella vaccines and tetanus, diphtheria and polio vaccines at the time of joining the study and at the end of the follow-up period. We saw a percentage increase of 363% for participants fully vaccinated for measles, mumps, rubella vaccines and a percentage increase of 86% for participants fully vaccinated for tetanus, diphtheria and polio vaccines at the end of follow-up, following implementation of our standardised study tool (Table 1).

Of the 52 participants referred for MMR (including one who was fully vaccinated at study start but received additional boosters through the study), 33 (64%) completed their required course (determined based on individual history and UK catch-up vaccination guidelines). Although our study was not powered to statistically compare the differences between sites, 33% [6] of referred participants in Site 1 had completed their required course of MMR by study end, compared to 84% [27] in Site 2 (Table 1).

51 participants were referred for Td/IPV vaccination (including 3 who reported being fully vaccinated on study enrolment but were referred for additional boosters). By the end of the follow-up period, 6 (12%) participants had completed their required course of Td/IPV, 40 (78%) had not, and 5 (10%) were unclear (due to limitations in the recording of their initial vaccination history data however they may have completed their course) (Table 1). There were again differences noted between sites (Table S4).

2 (4%) participants were eligible to be referred for MenACWY vaccine and 2 (4%) for HPV vaccination based on age. Both eligible participants were referred for MenACWY vaccine and one participant (50%) received one dose. 1 (50%) of the two eligible participants was referred for a HPV vaccine but did not receive it (Table S4).

### Qualitative findings

Through our FGDs with practice staff, we identified 7 key barriers to delivering catch-up vaccination in primary care (Figure 4). Staff offered several suggestions to strengthen delivery of catch-up vaccination in primary care, which we grouped into 7 main areas for improvement and support (Box 2). Suggested levels of responsibility (practice, system, and policy) for changes are also highlighted. The in-depth interviews with migrants highlighted a range of barriers to catch-up vaccinations, including rarely being offered catch-up vaccination upon arrival or upon presenting to a healthcare facility, as well as factors related to trust, safety and side effects, and preferences for natural immunity (full dataset reported elsewhere).

**Figure 4.**
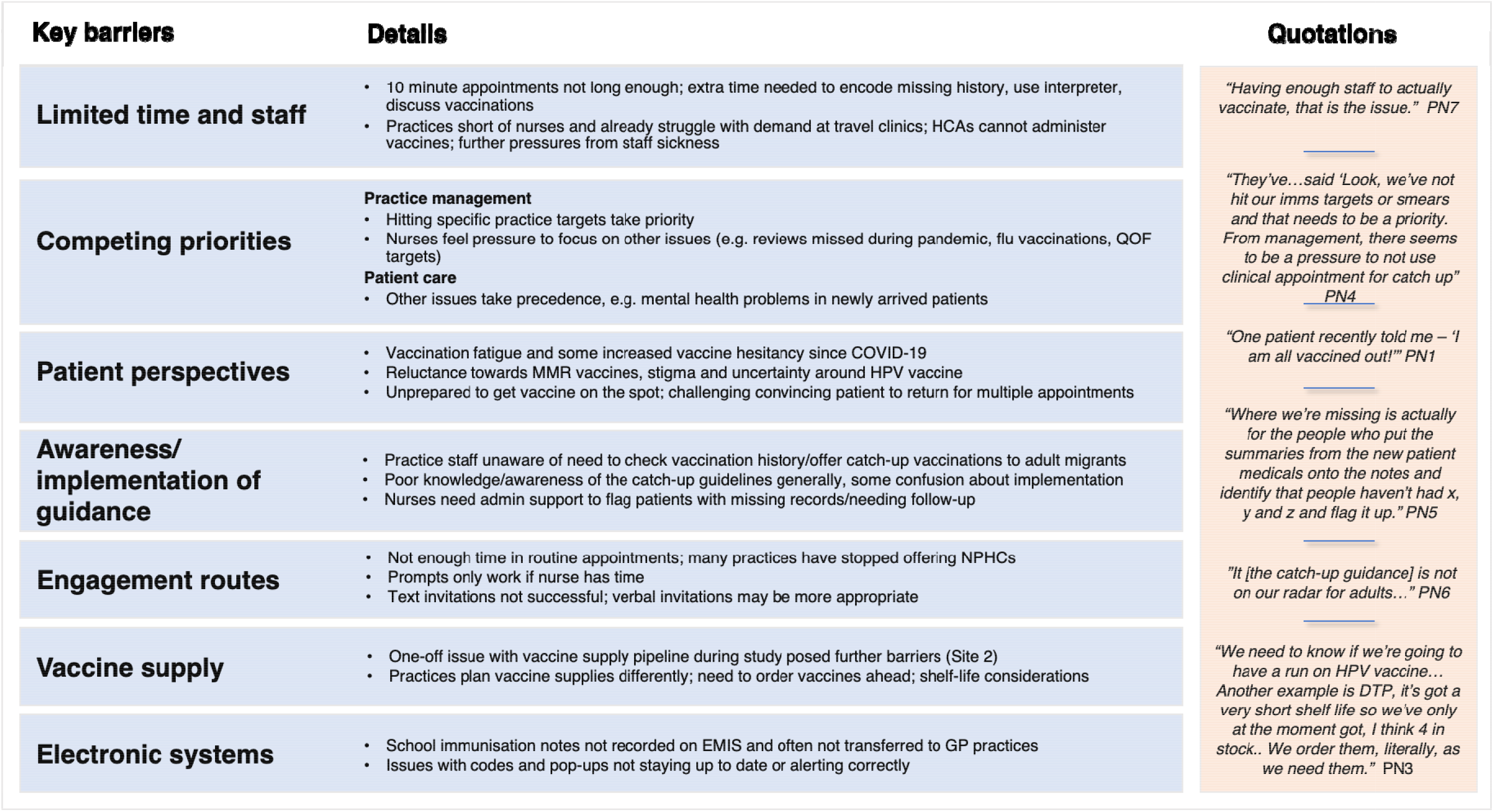
Key barriers to delivering catch-up vaccination in primary care with illustrative quotations, taken from 3 focus group discussions with 30 practice nurses/staff and 1 key informant interview with a practice nurse and assistant practice manager.

A case study of positive practice supporting catch-up vaccination of adult migrants in primary care based on findings from the key informant interview conducted with staff from Site 2 is highlighted in Figure 5.

**Figure 5.**
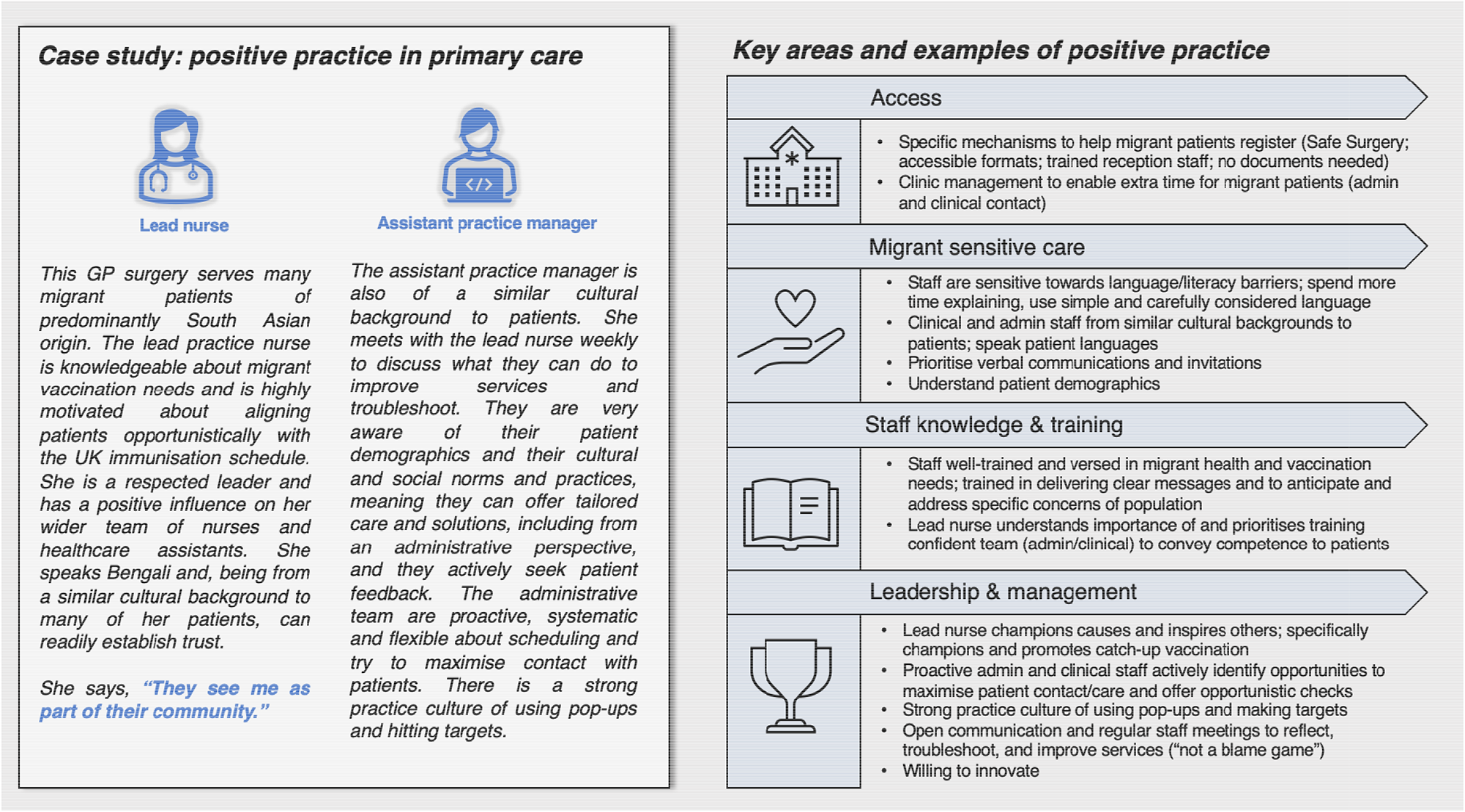
Case study of positive practice supporting catch-up vaccination in primary care, taken from focus group discussion with most successful participating GP surgery.

## Box 2.

**Improving catch-up vaccination: synthesis of staff suggestions and recommendations generated in FGDs**

**Strengthen and clarify existing guidance and data**

- Clearer, more specific guidance needed; decision trees for specific scenarios may help; involve staff in co-designing updated algorithm **(policy)**
- Unify European and UK catch-up guidelines to advance progress towards regional goals and ensure migrants are aligned with core schedule of vaccinations **(policy)**
- Introduce clear mandatory and statutory governance of a multi-disciplinary integrated immunisation leadership structure **(policy)**
- Routinely record migration status in electronic patient records and proactively check immunisation history of migrant patients attending primary care to identify those requiring catch-up vaccination. **(policy)**

**Explore and evaluate novel pathways, settings, approaches and funding mechanisms**

- Book new migrant patients in for immunisation reviews upon registering with a practice **(practice)**
- Trial novel engagement routes for catch-up vaccinations and innovative financing mechanisms to support delivery and implementation, e.g. designated clinics, longer and out-of-hours appointments, nurse-led community interventions, school- and work-based awareness raising and signposting **(practice/system/policy)**
- Identify and implement measures to reduce pressures in primary care and/or explore feasibility of shifting responsibility for catch-up vaccinations outside of primary care **(policy/system)**

**Set and use targets and incentives**

- Introduce government-backed catch-up vaccination targets and financial incentives in general practice; ensure immunisation targets and indicators are incorporated into integrated care strategies **(policy)**
- Use existing pop-ups and available data to identify potentially eligible patients opportunistically **(practice)**

**Facilitate and champion good practice**

- Normalise checking vaccination history and offering catch-up vaccination to migrant patients **(practice/system)**
- Identify staff champions to motivate, lead and inspire staff to meet targets **(practice/system)**
- Promote and establish mechanisms of shared working across primary care networks through infrastructure funding, IT systems, governance, shared flexible staffing, and HR agreements **(practice/system)**

**Provide training**

- Train clinical staff in motivational interviewing and conversational techniques to encourage vaccination uptake; provide training resources, e.g. speaking to vaccine hesitant patients and examples of answers to challenging questions; deliver training on migrant health needs and cultural competency **(practice/system)**
- Implement clear, appropriately-funded immunisation training pathways for healthcare professionals and continue to identify and expand primary care groups able to carry out immunisation training (e.g. pharmacists), enabling less experienced healthcare professionals to focus on delivery of less complex immunisation programmes and freeing experienced staff to focus on more complex areas, e.g. catch-up vaccination **(policy/system)**

**Tailor services**

- Understand patient demographics **(practice)**
- Use carefully considered wording and formats to invite patients and explain vaccination needs and opportunities **(practice)**
- Employ staff of similar cultural/linguistic backgrounds as patient population or create designated roles in primary care focused on community engagement and reducing barriers to services for marginalised groups **(practice/system/policy)**
- Utilise place-based partnerships (including NHS, local council, community and voluntary sector, local residents, service users, carers, representatives and community partners) to co-design and deliver integrated services to strengthen catch-up vaccination locally **(practice/system/policy)**

**Designate adequate funding and infrastructure**

- Expand funding and resources for routine childhood immunisations programmes to enable recall outside of the standard age groups for catch-up vaccination of adults and adolescents **(policy/system)**
- Ensure adequate funding for primary care nursing and community nursing workforce sustainability **(policy/system)**
- Reconcile issues with electronic/IT systems to ensure linkage between schools and GP immunisation records and wider system engagement **(practice/system)**
- Consider the roles of integrated care systems (ICSs) in delivering recommendations and improving quality, efficiency, equity, and outcomes **(system)**

## Discussion

We engaged a diverse group of migrants to the UK, the majority (>86%) of whom had incomplete or uncertain immunisation history for core vaccines in the UK immunisation programme, including MMR and Td/IPV, suggesting that migrants are an under-vaccinated group in the UK that could benefit from catch-up vaccination on arrival. Following implementation of our standardised data collection tool, the percentage of participants fully aligned with the UK schedule for measles, mumps, and rubella vaccines increased by 363%, and the percentage fully aligned for diphtheria, tetanus and polio vaccines increased by 86%. Most participants reported that they had not been previously offered catch-up vaccinations (MMR, Td/IPV and selectively, MenACWY and HPV) in the UK since their arrival, although half had been offered flu vaccine and some had been offered travel vaccines such as hepatitis A and typhoid, and most (93%) had received at least one dose of the COVID-19 vaccine.

Hence, even where migrants are registered with primary care, their catch-up vaccination needs are likely being overlooked, and catch-up guidelines are not being routinely implemented. Given that nearly all of our participants were assumed unimmunised for at least one, and typically many, VPDs the lack of significant predictors in our regression analyses are very much a result of the sample in the analysis. When catch-up vaccination was facilitated by study teams, 53 (93%) participants identified as under-immunised were referred. However, although 43 (81%) participants had received at least one dose of a required vaccine at follow-up, only 6 (12%) participants referred for Td/IPV and two thirds (33, 64%) of those referred for MMR had completed their required course and vaccination pathway at follow-up, suggesting there were a range of personal and environmental obstacles to migrants accessing vaccinations and multiple doses of vaccines that need to be better considered. Staff reported that there is rarely time in a routine appointment to engage migrants and offer catch-up vaccinations, and limited time to follow up patients to invite them for catch-up, which may require multiple doses over an extended timeframe – especially when trust, language and literacy barriers must also be overcome.

Although most patients attended a facilitated appointment with the practice nurse after being identified as requiring catch-up vaccination, drop out appears to have occurred for subsequent doses. There were also variations in interpretation of the guidelines, with some nurses recommending a full course of vaccinations regardless of previous history, and others recommending only the doses or boosters needed to complete a previously started course. This ambiguity in the catch-up vaccination guidelines/algorithm was also mentioned in the FGDs and is likely to be approached differently between practices and practitioners, indicating a need for clearer, more standardised procedures to support implementation in future. We found that Site 2 was more successful than Site 1 at starting participants on the catch-up vaccination pathway and facilitating further doses (100% started the pathway in Site 2 compared to 44.4% in Site 1, at end of follow-up period). This may be partly attributed to the positive influence of the lead nurse in Site 2, who championed catch-up vaccination among her team, was aware of the vaccination needs of her migrant patients and the catch-up guidelines and prioritised administering first doses immediately. This may also help to explain why 85% [27] of participants referred for MMR in Site 2 had completed their required course at the end of follow-up, compared to 33% [6] in Site 1. Some practices mentioned challenges with vaccine supply, and not being able to anticipate vaccine demand for adult patients as this is not currently factored into orders. If government targets around catch-up vaccination were introduced, staff may become more likely to prioritise catch-up vaccination processes routinely, and practices may be better able to anticipate vaccine supplies. We saw comparable (but notably low) proportions of participants starting the Td/IPV pathway between sites, but lower completion of the pathway in Site 2. This may be explained by a vaccine supply issue at Site 2 at the time of the study, which was highlighted by the lead nurse during the key informant interview. This meant that the practice prioritised MMR vaccinations while they waited for replenished Td/IPV vaccine stock.

Our findings showing low uptake for subsequent doses of catch-up vaccines also suggested that efforts may be needed to address some vaccination concerns, hesitancy, and fatigue (post-COVID-19) and build trust with certain migrant groups (28). This aligns with a recent systematic review which found acceptance barriers were mostly reported in Eastern European and Muslim migrant groups around HPV, measles and influenza vaccines (11). Another review highlighted sociodemographic and sociocultural barriers to HPV vaccination uptake (29). Providing primary care staff with specific training in motivational interviewing techniques to facilitate positive conversations around vaccination and encourage uptake is effective in some settings and for some vaccines. It has been made available to NHS Wales staff (30–32), and may be an important component of any future implementation efforts. Building trust through an ‘insider’ perspective (33), for example through a personalised approach and involving a trusted health professional from a shared community, may also be more effective than approaches involving a person perceived as an ‘outsider’ by the patient.

Our quantitative and qualitative findings combined point to several changes needed at policy, system, and practice levels to strengthen the delivery of catch-up vaccinations to under-vaccinated migrant groups. Notably, there is a need to strengthen and clarify existing guidance and promote catch-up vaccination of migrants in primary care. While catch-up vaccination guidelines for patients with no record or history of immunisation are straightforward, specific steps for those with partial immunity (e.g. conferred by single doses of vaccines or recollection of having a VPD such as measles as a child) are less clear and sometimes open to interpretation. Involving staff in co-designing an updated algorithm that addresses specific areas of confusion, adding decision aids and instructions for implementation, particularly to aid in identifying patients who may have missed routine vaccination, and introducing targets and training for catch-up vaccination in practices with high proportions of migrant patients are approaches that could be considered. Practices will likely prioritise hitting incentivised targets set in the current Quality and Outcomes Framework (QOF), which does not include catch-up vaccinations, particularly if they are overstretched or understaffed. It is unlikely that vaccination for migrants will be included in future versions of the QOF but financial and administrative support for vaccination in this group could be provided by Integrated Care Boards in England as part of local contractual arrangements for general practice. We found that identifying a staff champion who understands the guidance and can lead and motivate practice staff to deliver catch-up vaccination and meet targets was effective. Having practice staff from similar cultural and linguistic backgrounds as patients can also help instil patient trust (33) and improve the delivery of culturally competent care, while staff may be more invested in supporting causes and goals that benefit their community, including catch-up vaccination. Establishing peer or patient vaccine champion schemes or other designated community bridging roles in primary care, and using community-centred approaches, particularly in highly diverse, deprived, or underserved geographical areas, may also be an effective way to build trust, understand needs and encourage vaccine uptake and other positive behaviours within specific communities (34, 35), and could be done in partnership with local community and voluntary sector organisations and with Councils for Voluntary Service (CVS), NHS and local government support.

Even with stronger guidance and mechanisms to prompt staff to consider catch-up vaccination, barriers to delivery will remain due to intense pressures in primary care. Our data suggest that it may be more effective to explore and evaluate novel pathways, settings, and community-led or community-based outreach and interventions to deliver catch-up vaccination, and innovative financing mechanisms to support delivery and implementation. These could include, for example, offering catch-up vaccination through the New Patient Health Check or NHS Check in primary care, where there is more time to discuss preventative health care, conducting immunisation reviews for new patients, and using community settings, peer- or nurse-led interventions in the community, reducing or removing the burden from primary care. During the COVID-19 pandemic and the more recent London-based polio booster campaign, multiple innovations were seen in the delivery of vaccinations to marginalised groups, including offering incentives and flexible arrangements and infrastructure (e.g. out-of-hours clinics and alternative settings for vaccination (36–40). How these approaches could be used for delivering routine immunisations to migrants must be considered (41), ensuring the involvement and support of these communities in research and policy decisions. Working in partnership with community assets and networks to provide more localised and flexible approaches and outreach has been effective at facilitating attendance at NHS Health Checks (42) and should be explored. Ensuring migrants are involved in co-designing interventions that address their needs will also be vital (43).

Our findings align with much of the wider literature documenting the under-immunisation of migrants in Europe (1, 3, 44–46). We have uncovered similar barriers to delivering adult migrant catch-up vaccinations as studies done in the UK, Australia and Norway (9, 47, 48), including a lack of consistent guidelines, gaps in training and knowledge leading to missed opportunities by service providers, and perceptions that catch-up vaccination is time-consuming, difficult and resource intensive. Our study builds on this work through its practical efforts to pilot a novel digital tool to facilitate vaccine delivery in primary care, and its mixed-methods approach, which allowed for triangulation of data, a case study of good practice, and formulation of evidence-based recommendations. However, efforts are also needed to unify regional policy and increase the inclusion of adult migrants at the regional level. Evidence shows that policies and practices differ in European countries with respect to adult vaccination and the inclusion of adult migrants in vaccination programmes on arrival (4, 49, 50). For example, only 13/32 countries in the EU/EEA had policies in place to offer MMR to adult migrants (10 countries said they would charge fees). Addressing barriers at a regional level will be particularly important for meeting ECDC and WHO objectives to increase vaccine access and equity and ensure the integration of refugees and migrants in immunisation policies, service delivery and planning globally (14, 19, 51).

A strength of this study was its novel approach to assess under-vaccination and align migrant patients with the UK immunisation schedule, reducing their risk of contracting VPDs or experiencing ill health and closing immunisation gaps. Conducting this study during the COVID-19 pandemic, however, was challenging, with resource constraints and competing priorities in primary care posing additional complexities to implementation, as well as the need for modifications to our recruitment and care pathways in some participating GP practices, a much longer study period than we anticipated with low recruitment, and a shortened follow-up period in Site 2. However, these challenges reflect realities of primary care and led to valuable findings from our pilot study that can be used to inform future strategies and implementation. We also had major challenges recruiting patients in some practices, partly due to the poor recording of migrant status in electronic medical records, making it difficult to identify our target population, and due to very high work loads of practice nurses and CRN staff for the duration of our study. Low recruitment may also be reflective of migrant patients’ ‘vaccine fatigue’, reluctance to attend health facilities during the COVID-19 pandemic, or heightened mistrust or anticipated stigma (52–54) which made them less willing to receive or discuss vaccinations or engage with our study. The small sample size makes it difficult to generalise findings or draw wider conclusions; however, it provides an accurate snapshot of the levels of under-vaccination of many migrants presenting to UK primary care and highlights the lack of effective pathways for offering catch-up vaccination to these groups via primary care, which we will build on in a future large-scale trial.

## Data Availability

All data produced in the present study are available upon reasonable request to the authors

## Author contributions

SH conceptualised the study with input from ACF, AM and YC. AFC and SH wrote the ethics application and AFC and LG set up the study and analysed the data, with support from SH, JY, and DF. DF recruited patients. AD and AFC led the qualitative data component. AFC wrote the first draft with SH and all authors reviewed and commented on results and then a final version.

## Acknowledgments

We are grateful to and thank our participants who actively participated in the study, our study partners, participating GP practices, and healthcare staff. Thanks to Sobhash Jhuree and Mehwash Gondal, Central and North West London NHS Foundation Trust, Barnet Federated GPs Research Network, NHS North Central London Research Network, North Thames Clinical Research Network, and our St George’s, PPIE Project Advisory Board.

