## Supplemental tables for "Driving uptake of missed routine vaccines in adolescent and adult migrants: a prospective observational mixed-methods pilot study of catch-up vaccination in UK general practice"

**Main files (tables)**

**Table 1. Participants assumed un-immunised and fully vaccinated in line with UK schedule for measles, mumps, rubella, tetanus, diphtheria, and polio at time of joining study and at study end (N=57).**

| **Diseases** | **At time of joining study** | | **At study end** | | **Percentage increase in fully vaccinated participants by study end (%)** |
| --- | --- | --- | --- | --- | --- |
|  | Assumed unimmunised, n (%) | Fully vaccinated in line with UK schedule, n (%) | Assumed unimmunised, n (%) | Fully vaccinated in line with UK schedule, n (%) |  |
| Measles, mumps and rubella | 49 (86%) | 8 (14%) | 20 (35%) | 37 (65%) | 363 |
| Tetanus, diphtheria and  polio | 50 (88%) | 7 (12%) | 44^*^ (77%) | 13^*^ (23%) | 86 |

Participants were asked about their history of receiving combined and single vaccines. *Assumed unimmunised* = uncertain, missing, or incomplete vaccination history. *Fully vaccinated at study end* includes those who were fully vaccinated at time of joining study plus those who became fully vaccinated by receiving catch-up vaccination.

^*^Up to five more participants may have been fully vaccinated at study end (and therefore not ‘assumed unimmunised’) however not possible to report with confidence due to method of collecting data, where history of 1 or 2 previous doses of Td/IPV were coded as one variable.

**Supplementary files (tables)**

**Table S1. Socio-demographic characteristics of study population, by site and combined.**

| **Characteristic** | **Site 1 (Barnet, North London)** | **Site 2 (Tower Hamlets, East London)** | **Combined study population** |
| --- | --- | --- | --- |
| **Number of patients recruited** | 22 | 35 | 57 |
| **Number of GP practices that patients were recruited from** | 6 | 1 | 7 |
| **Mean age (SD, range)** | 41 (9.2, 22-63) | 41 (5.9, 24-51) | 41 (7.2, 22-63) |
| **Mean years in the UK^‡^ (SD, range)** | 13 (9.9, 0.5-40) | 9 (7.5, 0.1-33) | 11 (9.1, 0.1-40) |
| **Sex, n(%)** | | | |
| Female | 16 (72.7) | 19 (54.3) | 36 (62.1) |
| Male | 6 (27.3) | 16 (45.7) | 22 (37.9) |
| **Place of birth^*^, n(%)** | | | |
| UK | - | - | - |
| Europe (excluding UK) | 5 (22.7) | 0 | 5 (8.8) |
| Africa | 4 (18.2) | 0 | 4 (7.0) |
| Latin America/Caribbean | 2 (9.1) | 3 (8.6) | 5 (8.8) |
| Asia | 11 (50.0) | 32 (91.4) | 43 (75.4) |
| **Region lived immediately prior to UK, n (%)** | | | |
| Eastern Europe (Lithuania, Romania, Russia) | 4 (18.2) | 0 | 4 (7.0) |
| Africa (Algeria, Kenya, Nigeria, South Africa) | 3 (13.6) | 0 | 3 (5.3) |
| South America (Brazil, Chile) | 3 (13.6) | 3 (8.6) | 6 (10.5) |
| Asia (Bangladesh, China, Hong Kong, India, Iran, Japan, Philippines, Thailand) | 12 (54.6) | 31 (88.6) | 43 (75.4) |
| Missing | 0 | 1 (2.9) | 1 (1.8) |
| **Reason for migrating to UK^†^, n(%)** | | | |
| Economic reason | 11 (50.0) | - | - |
| To join or accompany family | 3 (13.6) | - | - |
| To study | 3 (13.6) | - | - |
| Forced migration | 4 (18.2) | - | - |
| No response | 1 (4.6) | - | - |
| **Occupation^†^, n(%)** |  |  |  |
| Lower-skilled | 6 (27.3) | - | - |
| Higher-skilled | 14 (63.6) | - | - |
| Student | 1 (4.6) | - | - |
| Not working | 1 (4.6) | - | - |
| **Had a vaccination card** | | | |
| Yes | 7 (31.8) | 2 (5.7) | 9 (15.8) |
| No | 15 (68.2) | 33 (94.3) | 48 (84.2) |

**^*^**Frequencies of birth countries not shown due to confidentiality but included: Algeria, Bangladesh, Brazil, Chile, China, Hong Kong, India, Iran, Japan, Kenya, Lithuania, Nigeria, Philippines, Romania, Russia, South Africa, Thailand. Countries grouped into regions according to United Nations geoscheme. Occupations organised according to official ONS Labour Force Survey categories.

**^‡^** Combined denominator n=40.

**^†^** Data not collected from Site 2.

**Table S2. Study participants’ vaccination history at time of recruitment.**

|  | **Site 1: Barnet, North London (N=22)** | **Site 2: Tower Hamlets, East London (N=35)** | **Combined study population (N=57)** |
| --- | --- | --- | --- |
| **History of vaccinations ever received in any country, n (%)** | | | |
| **Any measles-containing vaccine** |  |  |  |
| Yes | 7 (31.8) | 3 (8.6) | 10 (17.5) |
| No/DK | 15 (68.2) | 32 (91.4) | 47 (82.5) |
| **Any polio-containing vaccine** |  |  |  |
| Yes | 16 (72.7) | 14 (40.0) | 30 (52.6) |
| No/DK | 6 (27.3) | 21 (60.0) | 27 (47.4) |
| **MMR (combined vaccine)** |  |  |  |
| 2 doses | 5 (22.7) | 3 (8.6) | 8 (14.0) |
| 1 dose | 2 (9.1) | 0 | 2 (3.5) |
| 0 doses / don’t know (DK) | 15 (68.2) | 32 (91.4) | 47 (82.5) |
| **Td/IPV (combined vaccine)** |  |  |  |
| 3 doses | 4 (18.2) | 3 (8.6) | 7 (12.3) |
| 1-2 doses | 6 (27.3) | 11 (31.4) | 17 (29.8) |
| 0 doses/DK | 12 (54.6) | 21(60.0) | 33 (57.9) |
| **Measles (single vaccine)** |  |  |  |
| Yes | 6 (27.3) | 3 (8.6) | 9 (15.8) |
| No | 10 (45.5) | 32 (91.4) | 42 (73.7) |
| DK | 6 (27.3) | 0 | 6 (10.5) |
| **Mumps (single vaccine)** |  |  |  |
| Yes | 7 (31.8) | 3 (8.6) | 10 (17.5) |
| No | 10 (45.5) | 32 (91.4) | 42 (73.7) |
| DK | 5 (22.7) | 0 | 5 (8.8) |
| **Rubella (single vaccine)** |  |  |  |
| Yes | 7 (31.8) | 0 | 7 (12.3) |
| No | 9 (40.9) | 35 (100.0) | 44 (77.2) |
| DK | 6 (27.3) | 0 | 6 (10.5) |
| **Polio (single vaccine)** |  |  |  |
| Yes | 15 (68.2) | 12 (34.3) | 27 (47.4) |
| No | 7 (31.8) | 23 (65.7) | 30 (52.6) |
| DK | 0 | 0 | 0 |
| **Tetanus (single vaccine)** |  |  |  |
| Yes | 14 (63.6) | 13 (37.1) | 27 (47.4) |
| No | 6 (27.3) | 22 (62.9) | 28 (49.1) |
| DK | 2 (9.1) | 0 | 2 (3.5) |
| **TB vaccine/BCG** |  |  |  |
| Yes | 8 (26.4) | 0 | 8 (14.0) |
| No | 10 (45.5) | 35 (100.0) | 45 (79.0) |
| DK | 4 (18.2) | 0 | 4 (7.0) |
| **MenACWY vaccine (bacterial meningitis)** |  |  |  |
| Yes | 6 (27.3) | 5 (14.3) | 11 (19.3) |
| No | 13 (59.1) | 30 (85.7) | 43 (75.4) |
| DK | 3 (13.6) | 0 | 3 (5.3) |
| **Hepatitis A** **vaccine** |  |  |  |
| Yes | 6 (27.3) | 12 (34.3) | 18 (31.6) |
| No | 11 (50.0) | 23 (65.7) | 34 (59.7) |
| DK | 5 (22.7) | 0 | 5 (8.8) |
| **Hepatitis B** **vaccine** |  |  |  |
| Yes | 4 (18.2) | 1 (2.9) | 5 (8.8) |
| No | 14 (63.6) | 34 (97.1) | 48 (84.2) |
| DK | 4 (18.2) | 0 | 4 (7.0) |
| **Human Papillomavirus (HPV)** **vaccine** |  |  |  |
| Yes | 12 (54.6) | 2 (5.7) | 14 (24.6) |
| No | 8 (26.4) | 33 (94.3) | 41 (71.9) |
| DK | 2 (9.1) | 0 | 2 (3.5) |
| **COVID-19 vaccine** |  |  |  |
| Yes | 20 (90.9) | 33 (94.3) | 53 (93.0) |
| No | 2 (9.1) | 2 (5.7) | 4 (7.0) |
| **History of vaccinations ever offered or received in the UK prior to this study** | | | |
| **Ever offered any vaccines in the UK (by any service) n (%)** |  |  |  |
| Yes | 22 (100.0) | 35 (100.0) | 57 (100.0) |
| No | 0 | 0 | 0 |
| **Received offered vaccines, n (%)** |  |  |  |
| Yes | 18 (81.8) | 32 (91.4) | 50 (87.7) |
| No | 1 (4.6) | 1 (2.9) | 2 (3.5) |
| Not applicable | 2 (9.1) | 0 | 2 (3.5) |
| Not specified | 1 (4.6) | 2 (5.7) | 3 (5.3) |
| **Vaccines offered, n(%)** |  |  |  |
| MMR | 1 (4.6) | 2 (5.7) | 3 (5.3) |
| Td/IPV or DTP | 1 (4.6) | 4 (11.4) | 5 (8.8) |
| MenACWY | 0 | 2 (5.7) | 2 (3.5) |
| HPV | 0 | 0 | 0 |
| Influenza | 6 (27.3) | 23 (65.7) | 29 (50.9) |
| COVID-19 | 22 (100.0) | 35 (100.0) | 57 (100.0) |
| Hepatitis B | 3 (13.6) | 1 (2.9) | 4 (7.0) |
| Hepatitis A | 2 (9.1) | 10 (28.6) | 12 (21.1) |
| Typhoid | 2 (9.1) | 8 (22.9) | 10 (17.5) |
| Pneumococcal vaccine | 0 | 4 (11.4) | 4 (7.0) |
| Other (varicella, pertussis, diphtheria, tetanus, unspecified travel vaccines) | 5 (22.7) | 3 (8.6) | 8 (14.0) |

**Table S3. Participants’ history of VPDs at time of study recruitment.**

|  | **Site 1: Barnet, North London (N=22)** | **Site 2: Tower Hamlets, East London (N=35)** | **Combined study population (N=57)** |
| --- | --- | --- | --- |
| **Recalled having a vaccine preventable disease (not including COVID-19)^†^, n (%)** |  |  |  |
| Yes | 12 (54.6) | - | - |
| No | 8 (36.4) | - | - |
| DK | 2 (9.1) | - | - |
| **Reported having had 2 or more vaccine preventable diseases (not including COVID-19)^†^, n(%)** |  |  |  |
| Yes | 3 (13.6) |  |  |
| No/DK | 19 (86.4) |  |  |
| **Historical vaccine preventable disease cases reported, n(%)** |  |  |  |
| Measles | 2 (9.1) | - | - |
| Rubella | 1 (4.5) | - | - |
| Active TB | 1 (4.5) | - | - |
| Bacterial meningitis | 1 (4.5) | - | - |
| Pertussis | 2 (9.1) | - | - |
| Hepatitis A | 1 (4.5) | - | - |
| Hepatitis B | 1 (4.5) | - | - |
| HPV | 5 (22.7) | - | - |
| COVID-19 | 17 (77.3) | 33 (94.3) | 50 (87.7) |

^†^Data on history of vaccine preventable disease were only collected from Site 1 and included: measles, mumps, rubella, diphtheria, pertussis, polio, tetanus, active TB, bacterial meningitis, hepatitis A, hepatitis B, HPV. History of COVID-19 was counted separately.

**Table S4. Catch-up vaccination referral and uptake as part of the study.**

|  | **Site 1: Barnet, North London (N=22)** | **Site 2: Tower Hamlets, East London (N=35)** | **Combined study population (N=57)** |
| --- | --- | --- | --- |
| **Patients referred for any catch-up vaccination, n (%)** | | | |
| Yes | 18 (81.8) | 35 (100.0) | 53 (93.0) |
| No | 4 (18.2) | 0 | 4 (7.0) |
| **Completed required course of Td/IPV, n (%)** | N=19 referred | N=32 referred | N=51 referred* |
| Yes | 6 (31.6) | 0 | 6 (11.8) |
| No | 12 (63.2) | 28 (87.5) | 40 (78.4) |
| Not clear | 1 (5.3) | 4 (12.5) | 5 (9.8) |
| **Completed required course of MMR, n (%)** | N=18 referred | N=34 referred | N=52 referred^^^ |
| Yes | 6 (33.3) | 27 (84.4) | 33 (63.5) |
| No | 12 (66.7) | 7 (21.9) | 19 (36.5) |
| Not clear | 0 | 0 | 0 |
| **Started required course for MenACWY, n (%)** | N=1 referred | N=1 referred | N=2 referred |
| Yes – 1 dose | 0 | 1 (100.0) | 1 (50.0) |
| No | 1 (100.0) | 0 | 1 (50.0) |
| **Started required course for HPV, n (%)** | N=1 referred | N=0 referred | N=1 referred |
| Yes | 0 | 0 | 0 |
| No | 1 (100.0) | 1 (100.0) | 1 (100.0) |
| **Reason given for not attending catch-up vaccination appointment, n (%)** | | | |
| Did not respond to invitation | 5/10 (50.0) | - | 5/10 (50.0) |
| Wanted to check vaccination records first | 1/10 (10.0) | - | 1/10 (10.0) |
| Not followed up after recruitment | 2/10 (20.0) | - | 2/10 (20.0) |
| Reason not given | 2/10 (20.0) | - | 2/10 (20.0) |

*One patient who reported being fully vaccinated (3 doses) for Td/IPV received 2 further doses of Td/IPV through the study and is included here.

^^^ 3 participants who reported being fully vaccinated (2 doses) for MMR received boosters through the study and are included here.

Follow-up period was up to 14 months in Site 1 (median 12 months) and 6 months in Site 2.

**Table S5. Logistic regression analyses showing factors associated with un-vaccination for polio (zero doses of single or combined vaccines).**

| Variable | Sample size | No. unvaccinated | Unadjusted OR (95% CI) | P value | Adjusted OR (95% CI) | P value |
| --- | --- | --- | --- | --- | --- | --- |
| Outcome: Unvaccinated for polio vaccine | | | | | | |
| Age (years) | 55 | 25 | 1.02 (0.94-1.11) | 0.61 | **1.21 (1.01-1.47)** | **<0.05 (0.042)** |
| Sex |  |  |  |  |  |  |
| Female | 35 | 19 | 1 (0.34-2.95) | 1.00 | 2.33 (0.33-16.45) | 0.40 |
| Male | 22 | 11 | 1 |  |  |  |
| Birth region |  |  |  |  |  |  |
| Europe* | 5 | 2 | 1 |  | 1 |  |
| Africa | 4 | 1 | 1 (empty) |  | 1 (empty) |  |
| Latin America/ Caribbean | 5 | 3 | 1 (0.80-12.56) | 1.00 | 0.50 (0.02-13.03) | 0.67 |
| Asia | 43 | 24 | 1.5 (0.23-9.92) | 0.67 | 1.82 (0.13-24.72) | 0.65 |
| Region prior to UK |  |  |  |  |  |  |
| Europe* | 4 | 2 | 1 |  |  |  |
| Africa | 3 | 0 | 1 (empty) | 0.55 |  |  |
| Latin America/ Caribbean | 6 | 3 | 0.5 (0.04-6.68) | 0.60 |  |  |
| Asia | 43 | 25 | 1.05 (0.13-8.18) | 0.96 |  |  |
| Years in UK | 40 | 20 | 0.82 (0.70-0.96) | <0.05 (0.016) | **0.75 (0.60-0.94)** | **<0.05 (0.013)** |
| Study site |  |  |  |  |  |  |
| Site 1* | 22 | 7 | 1 |  |  |  |
| Site 2 | 35 | 23 | 6.00 (1.66-21.74) | <0.006 |  |  |

OR, odds ratio; aOR, adjusted odds ratio; CI, confidence interval. Unadjusted and adjusted odds ratios calculated using generalised estimating equations logistic regression. *Comparison group. Significant values (p<0.05) shown in bold. Outcomes were binary (not vaccinated versus received at least 1 dose). Country prior to UK was removed from final model to reduce collinearity with birth region; study site was removed to reduce collinearity.

**Table S6. Comparisons of distributions of sociodemographic values between study sites (Site 1, N=22; Site 2, N=35), performed using Pearson’s Chi-squared test and unpaired t-tests.**

| **Variable** | **Test value** | **df** | **p value** |
| --- | --- | --- | --- |
| Age | 0.2008 | 55 | 0.84 |
| Years in UK | 1.6805 | 38 | 0.10 |
| Sex | 1.9385 | 1 | 0.16 |
| Region of birth | 17.3958 | 3 | **0.01** |
| Region lived immediately prior to UK | 13.4411 | 3 | **<0.01 (0.004)** |
| Had vaccination card | 6.9230 | 1 | **<0.01 (0.009)** |

X^2^, Pearson’s Chi-squared test; df, degrees of freedom. Pearson’s Chi-squared test performed on categorical and ordinal variables; unpaired t-tests performed on continuous variables (age and years in UK). Significant values (p<0.05) shown in bold.

**Table S7. COVID-19 vaccination offers and uptake in the UK.**

|  | **Site 1: Barnet, North London (N=22)** | **Site 2: Tower Hamlets, East London (N=35)** | **Combined study population (N=57)** |
| --- | --- | --- | --- |
| **Invited to receive COVID-19 vaccine in UK, n (%)** |  |  |  |
| Yes | 22 (100.0) | 35 (100.0) | 57 (100.0) |
| No | 0 | 0 | 0 |
| **Accepted COVID-19 vaccine offer, n(%)** |  |  |  |
| Yes | 20 (90.9) | 33 (94.3) | 53 (93.0) |
| No | 2 (9.1) | 2 (5.7) | 4 (7.0) |
| **COVID-19 vaccine – number of doses received** |  |  |  |
| 4 doses | 0 | 0 | 0 |
| 3 doses | 4 (18.2) | 27 (77.1) | 31 (54.4) |
| 2 doses | 14 (63.6) | 6 (17.1) | 20 (35.1) |
| 1 dose | 2 (9.1) | 0 | 2 (3.5) |
| 0 doses | 2 (9.1) | 2 (5.7) | 4 (7.0) |
| No data | 0 | 0 | 0 |
| **Reason for not accepting COVID-19 vaccine offer, n(%)** |  |  |  |
| Medical reason | 1 (50.0) | 0 | 1 (25.0) |
| Not specified | 1 (50.0) | 2 (100.0) | 3 (75.0) |
